# Tissue Stress from Laparoscopic Grasper Use and Bowel Injury in Humans: Establishing Intraoperative Force Boundaries

**DOI:** 10.1101/2021.02.19.21252109

**Authors:** Amanda Farah Khan, Matthew Kenneth MacDonald, Catherine Streutker, Corwyn Rowsell, James Drake, Teodor Grantcharov

## Abstract

**Background:** Inappropriate force in laparoscopic surgery can lead to inadvertent tissue injury. It is currently unknown however at what magnitude of compressive stress trauma occurs in gastrointestinal tissues.

**Methods:** This study included 10 small bowel and 10 colon samples. Each was compressed with pressures ranging from 100 kPa to 600 kPa by a novel device to induce compressive “grasps” to simulate those of a laparoscopic grasper. Experimentation was performed *ex-vivo, in-vitro*. Grasp conditions of 0 to 600 kPa for a duration of 10 seconds were utilized. Two pathologists who were blinded to all study conditions, performed a histological analysis of the tissues. Patients were eligible if their surgery procured healthy tissue margins for experimentation (a convenience sample). 26 patient samples were procured; six samples were unusable. 10 colon and 10 small bowel samples were tested for a total of 120 experimental cases. No patients withdrew their consent. Two metrics of damage were quantified: an intestinal layer thickness calculation where the serosa layer was measured in the area of compression and compared to a local control and a histological scoring scale for tissue trauma.

**Results:** Small bowel (10), M:F was 7:3, average age was 54.3 years. Colon (10), M:F was 1:1, average age was 65.2 years. All 20 patients experienced a significant difference (p<0.05) in serosal thickness post-compression at both 500 and 600 kPa for both tissue types. A logistic regression analysis with a sensitivity of 100% and a specificity of 84.6% on a test set of data predicts a safety threshold of 329-330 kPa.

**Conclusion:** A threshold was discovered that corresponded to both significant serosal thickness change and a positive histological trauma score rating. This “force limit” could be used in novel sensorized laparoscopic tools to avoid intraoperative tissue injury.

## Introduction

Laparoscopic graspers are used extensively in minimally invasive surgery (MIS), primarily to lift and mobilize delicate anatomical tissues for better visualization and access. With MIS procedures becoming increasingly common in general surgery, gynecology and urology, the problems that arise with the use of such tools must be carefully considered [1]. For example, serious iatrogenic complications from the improper use of laparoscopic graspers in bowel surgery include: bowel perforation, serosal tears and post-operative adhesion formation [2]. Bowel perforation is an especially severe complication because it is associated with a high morbidity and mortality rate (as high as 3.6%) but its incidence is entirely due to intraoperative error from the misuse of graspers [3], [4]. Tang et al. studied the removal of the gallbladder, a routine gastrointestinal laparoscopic procedure. They found that the majority of intraoperative errors occurred from grasper use and that 11.3% of consequential and 19% of inconsequential grasper-related injuries were due to excessive force [5]. Other delicate tissues that are susceptible to grasper injuries include the bile duct, ureters, fallopian tubes and spleen.

Careful evaluation of these statistics is especially pertinent in light of the fact that 100 patients a day die from iatrogenic injuries in United States (US) hospitals, with 40% of these injuries occurring in the operating room [6], [7]. In Canada, Baker et al., studied adverse events occurring in hospitals across five different provinces. They found that 7.5% of all patients admitted to acute care hospitals experienced one or more adverse event(s), with 51.4% of all adverse events arising from surgery. They judged that 36.9% of these adverse events were *highly preventable* [8].

Unfortunately, there has been a dearth of studies that attempt to quantify the interaction of the grasper-tissue interface at a histological level to quantify which load forces tissue injuries occur at. This topic is particularly important to explore, because researchers have found that the handle and tip forces in laparoscopic graspers differ significantly from conventional graspers used in “open-approach” surgeries, which can lead to inappropriate force magnitudes and tissue damage [9], [10]. Stress thresholds need to be established to limit the likelihood of this from occurring.

### The Complex Mechanical Response of Tissue to Compression

It is challenging to accurately quantify and model biological soft tissues’ multifaceted and complex behavior in response to the compressive force exerted by laparoscopic graspers. The mechanical response of tissue is based on two factors: a) the inherent mechanical properties of that tissue and b) the environmental loading characteristics it is subjected to. The small and large bowel are composed of multiple tissue layers. Within each layer, different fibers are distributed according to specific spatial orientations, which creates a strongly anisotropic configuration where measured properties varies along its different axes [11]. For example, colonic tissues have been shown to exhibit a non-linear viscoelastic rate-dependent response under compression due to its inhomogeneity [12].

### Grasper Jaw Geometry and Stress on Tissues

Laparoscopic graspers have jaws traditionally made from stainless steel due to its durability and ease of sterilization. The main disadvantage of using metal is that metal is a much stiffer material than delicate gastrointestinal tissues and as such, compressing tissue with metal graspers can cause damage at the cellular level (such as mechanical destruction of the cell membrane or nucleus) or tissue level (such as rupture of muscle fibers or ischemia from the destruction of blood vessels) [13]. Graspers also come in a variety of jaw geometries and teeth profiles such as straight or flared, fenestrated with waves or solid and single or dual action, which contribute both to their function and damage potential. When designing a new grasper, consideration for jaw profile must be first and foremost because the jaw must maximize tissue grip to avoid slippage but it also must not damage tissue by creating areas of stress concentration. For example, when a grasper’s jaw is serrated, increasing the size of the teeth will help prevent slippage but also causes more damage to tissue [14]. Rounding the edge of the graspers can reduce high stress concentrations at the tip [15]. Cheng and Hannaford further investigated this relationship and created both a 2D and 3D finite element analysis (FEA) study of calculated von Mises stress distributions under compression loads in a grasper and liver tissue model [16]. They found that in the 2D plane strain model, that 80% of the stress in the area directly beneath the grasper was over 300 kPa, which is over the damage limit they elucidated in their previous work of 240 kPa for liver tissue [17]. Injury to tissue can also increase over time, even after the initial stress event is over. This happens when a core amount of tissue experiences necrosis from the initial pressure event and then stiffens. This stiffening may induce adjoining cells to have an increase in osmotic pressure, a decrease in efficiency of cellular mechanisms of repair and subsequent necrosis of the adjoining tissue. Nagel et al., explored the effects of cellular stiffening on tissue damage and found that stiffening significantly contributed to both the pressure induced damage amount and the rate of damage progression [18].

### Is Porcine Tissue an Accurate Surrogate for Human Tissues?

Previous studies exploring grasper jaw and tissue interactions have mostly centered on porcine tissue studies. This is due to the vast logistical and ethical challenges involved in human tissue experimentation and the previous assumption that porcine tissues are close enough to human tissues to be a surrogate model. Christensen et al.’s work with porcine and human bowel tissues casts doubt on this assumption however, as they found that human tissues were stronger, stiffer and less compliant than porcine tissue. Porcine tissue was able to stretch almost twice as much as human bowel tissue (with an elastic modulus of 1.83 MPa and 5.18 respectively), while human bowel tissue had a higher ultimate average strength (0.58 MPa compared to 0.87 for human tissues) [19]. Heijnsdijk et al. also found that the inter-individual variability in perforation forces is quite large and that bowel strength could differ by a factor of two between patients [20]. Because of this large variation in bowel strength, surgeons must be acutely aware that forces that can be safely applied to one patient may cause a perforation in another. Both groups’ data suggests that porcine tissue does not accurately model human bowel tissue’s mechanical properties and that new experimentation with human tissues must occur to establish accurate tool-tissue force limits.

### Establishing Safe Tissue Force Boundaries in Humans with a Histopathological Analysis

This study aims to address this concern by investigating the relationship between grasper jaw forces and human small and large bowel (colon) tissues. These tissue types were specifically chosen for two reasons: first, they are the most clinically relevant in relation to repeated grasp injury and second, bowel is one of the most delicate tissues in the human body. van der Voort et al., found that the overall incidence of laparoscopy-induced bowel injury was 0.36%. The small intestine was most frequently injured (55.8%), followed by the large intestine (38.6%) [3]. Schwartz et al. found an even higher incidence of 0.65% with blunt injury (23.1%) being the most common etiology of damage [21].

To the authors’ knowledge, this will be the first study to investigate the upper limit of force by laparoscopic graspers in human tissues with a histological analysis of cellular damage. A histological model was chosen to objectively and quantifiably understand how the intestinal tissue structure is microscopically affected as a result of mechanical loading. This data will be important as we move into an age of “smart surgical tools” that can quantify tool-tissue force interactions. This data must also be incorporated into the training of new surgical residents. While the McGill Inanimate System for Training and Evaluation of Laparoscopic Skills (MISTELS) is widely used to teach and assess laparoscopic ability, assessment metrics do not currently take into account tool-tissue force interactions [22]. The lack of evaluation of the physical forces exerted on tissues misses a crucial dimension of surgical skill. Quantifying how surgical residents interact with delicate tissues should not just look at task time completion or economy of movement. A laparoscopic grasper with an integrated force sensor should be used so that force metrics can be evaluated as well.

## Materials and Methods

### Study Design and Experimental Protocol

We previously created a prototype device, the SimpleCAT (simple crush apparatus for tissue) to produce discrete grasp forces on human gastrointestinal tissue to test feasibility and workflow for human tissue experimentation [23]. This current paper aims to build upon the work of Chandler et al. in defining an upper force limit for human small and large bowel tissue with a more sophisticated and accurate device than our previous work [24]. To this end, a new custom device called the Precision Crush Apparatus for Tissue (PrecisionCAT) was created (Figure 1a). A study to test the device was approved by the Research Ethics Office of St. Michael’s Hospital (REB #15-299) in Toronto, Ontario. Patients were consented by their operating surgeon, using a standard surgical consent form, which at our institution, includes a provision for using excess surgical tissues (not needed for surgical pathology) for research purposes. Patients were given an information form as well, that included a contact number if they chose to withdraw their sample from the study at any time post-operation. All methods were performed in accordance with the relevant guidelines and regulations of our institution and ethics office.

Each patient had six 1 cm x 1 cm tissue samples cut from their usable tissue. Each sample was loaded onto the grasper plate of the PrecisionCAT serosa side up on a small cellulose-fiber sheet to avoid tissue slippage, and one at a time, a discrete force was exerted on it ranging from 0 to 11.8 N (0 to 600 kPa). Specifically forces of 2 N, 3.9 N, 5.9 N, 7.8 N, 9.8 N and 11.8 N were used, or using our precision pin plate with known surface area of 19.6mm^2^, calculated pressures of 0 kPa, 100 kPa, 200 kPa, 300 kPa, 400 kPa, 500 kPa and 600 kPa. Each simulated grasp was for a duration of 10 seconds. This is consistent with our previous protocol which used precision weights (0 g, 200 g, 400 g, 600 g, 800 g, 1000 g and 1200 g) and the same diameter of pin plate to generate equivalent force and pressure. These metrics were chosen to be consistent with the mean grasp force and 95% of grasp time elucidated by the Blue DRAGON system for laparoscopic tasks (8.52 N ± 2.77 N and 8.86 s ± 7.06 s), Zhou et al.’s tribology studies (0-16 N with significant tissue damage achieved past 13 N) and Chandler et al.’s protocol (0 - 300 kPa grasps with 10 s duration) [4], [25], [26]. The order in which tissues were compressed with each amount of force was randomized so that the pathologists did not know the tissue’s loading condition. After each tissue sample was compressed, it was cut in halfand processed for histology. On average, from the time the sample was removed from the patient to the end of the full experimental protocol, the research team took only 20 minutes, to maximally preserve tissue integrity. When analyzing the slides, the two pathologists were again blinded to the amount of force each tissue was subjected to and slides were analyzed out of order to prevent bias.

### Experimental Test Equipment

A test system (Figure 1a, Build of Materials in the Appendix) was developed based on Chandler et al.’s work, to apply mechanically controlled ‘grasps’ to tissue samples that are characteristic of those sustained intraoperatively. Key requirements were similar to Chandler’s: the test system should apply compressive grasps where the loading rate, peak stress and hold time are directly controlled and the grasper plate position precisely known. The ensuing system, named the Precision Crush Apparatus for Tissue (PrecisionCAT), uses a linear actuator with 0.1 µm resolution (LCA50-025-72-1F-3, SMAC Moving Coil Actuators, California, USA) to drive together two “grasp plates” (representing the grasper jaws) and thus compress a sample of target tissue. The actuator was controlled via the SMAC LAC-1 servo motor controller (SMAC Moving Coil Actuators, California, USA) and utilized force data from the load cell to regulate velocity and position. The load cell used was a high precision compression-link load cell (LCM-703-25, Omega Engineering, Connecticut, USA) coupled with a precision differential instrumentation amplifier (DMD-465, Omega Engineering, Connecticut, USA) to amplify and convert the load cell’s voltage to a larger voltage for digitization. Position and load data was synchronously recorded at 31 Hz. The grasp plates were rapid prototyped at 75 μm resolution using a Somos WaterShed XC 11122 (ZRapid SL600, Jiangsu, China). The plates’ geometry is not based on a specific grasper jaw with fenestrations, but instead, comprise of a smooth bottom test platform and the top, of a pin plate with an indentation tip, with the pin plate being driven closer to the test platform plate by the linear actuator (Figure 1b). The cylindrical pin was fabricated with a known diameter of 5 mm and surface area of 19.6 mm^2^ to allow for pressure to be calculated from force for a standardized measurement.

The geometry of the indentation tip and its size relative to the test material and test platform plate were important design considerations. A smooth, flat indentation tip with no fenestrations was used rather than actual grasper jaw geometry because a uniform circular flat tip ensures a constant and predictable contact profile between the material and the tip. This simplifies analysis as it avoids the difficulty of mapping applied pressure generated from a grasper jaw with fenestrations and a hinge mechanism. It also isolates strain effects on the tissue from the confounding effects of grasper geometry. Pressure would vary along the length of the jaw and depend on the mechanical advantage at the grasper linkage mechanism and would include areas of local stress at the peaks of the fenestrations. A large sample-to-tip dimension ratio also fulfills the half-space assumption used during analytical derivation for material testing [27].

The ability of the SimpleCAT to produce a consistent, even pressure on the loaded tissues was evaluated using Fujifilm Prescale film (Fujifilm, Tokyo, Japan). Prescale film can precisely measure pressure distribution and balance. The two-sheet system for Extreme Low Pressure was used (Prescale 4LW, R310 3M, 0.05 MPa). Both sheets are coated in a polyester base, with the top sheet impregnated with a color-developing material and the bottom coated with a micro-encapsulated color-forming chemical. When pressure is applied, the color microcapsules are broken and the color-developing material reacts to develop a red colour. The colour density and distribution corresponds to the uniformity of contact pressure. The SimpleCAT produced an even, consistent colour change on the Prescale film.

The full assembly schematics, .STL files and build of materials is available to use for free, via our Harvard Dataverse repository [28]. Existing Python code was adapted and expanded to create custom software to control the actuator motion and hold its position once a pre-defined load threshold from the load cell was reached [29]. This code is available to use freely on our Github repository [30].

### Biospecimen Tissue Preparation and Analysis

Small and large bowel tissues were chosen for this study based on data from our pilot project and the clinical significance of perforation or injury of these tissues [23]. Out of necessity, tissues included in this study were a convenience sample and were based on what operations were scheduled. All surgeons in the Division of General Surgery at St. Michael’s Hospital were enrolled in the study and when they had a scheduled operation that included a surgical pathology sample (with wide enough margins of normal tissue), our research team would be present in the operating room and acquire the sample as soon as it was removed from the body (Figure 1c). Small or large bowel was either removed laparoscopically or through an open surgical approach. Samples were kept in a fresh state rather than being stored in 10% buffered formalin to preserve cellular integrity and mechanical properties. Once removed, samples were immediately taken to an adjacent histology suite where the two pathologists, CS and CR would assess the tissue so that sections for testing were taken from tissue not needed for normal pathologic analysis. Segments of tissue that were inflamed, pathological or damaged by clips or sutures were excluded and only the normal, healthy tissue away from margins and lesional tissue were used. Six 1 cm x 1 cm tissue squares were cut from each sample and each loaded onto the PrecisionCAT serosa side up for experimentation. Full “tube” bowel sections were not used due to the amount of tissue available. Blue tissue marking dye (#1003-5 Blue, Davidson Marking System, Minnesota, USA) was used to coat the indentation tip of the grasp plate before the experiment was performed so that when the tip made contact with the tissue, the area of compression would be evident when preparing and viewing slides. Numerous dyes were tested such as India ink but this specific dye was found to be most reliably visualized under the microscope after slide processing. Once the experimental protocol was complete, the tested section was cut in half across the area of compression, fixed in formalin for normal histology processing and cut en-face to allow a full cross-sectional view across the area of compression, then regular tissue processing and staining took place, as follows: Tissues were fixed in 10% buffered formalin, processed and embedded in paraffin. Sections with a thickness of 4 microns were cut and tissues were mounted to glass slides. Careful consideration in regard to orientation needed to occur to ensure that sections that were parallel to the direction of the applied pressure was chosen, however in some cases the tissue twisted during processing/embedding and a flat transverse section was not possible. Sections were stained with hematoxylin and eosin (H&E) for typical visualization of cell morphology and structure. Once slide preparation was complete, they were scanned at 20x using a brightfield digital pathology scanner utilizing the time delay and integration (TDI) line scan method (Aperio AT Turbo, Leica Biosystems, Wetzlar, Germany). Slides were analyzed using digital slide viewing software (Aperio ImageScope, Leica Biosystems, Wetzlar, Germany) that included magnification, pan, zoom and distance measurement tools. Serosal thickness was specifically targeted for quantification because it is serosal disruption that is hypothesized to be the basis of adhesion formation. Peritoneal adhesions are found in up to 93% of patients post intra-abdominal surgery [31]. Adhesions can cause a number of significant clinical problems including bowel obstruction, chronic abdominal pain, infertility and organ tissue injury, leading to higher rates of morbidity and mortality post-surgery. It is hypothesized that trauma to serosal surfaces and the subsequent altered complex healing cascade can lead to permanent fibrin bridges that can form the basis of intestinal adhesion attachment to neighbouring structures such as other intestinal loops or the abdominal wall [31].

### Tissue Trauma Score and Serosa Thickness Calculations

Due to the experimental protocol being *ex-vivo*, normal markers of cellular injury such as increased granulocyte recruitment (neutrophils and eosinophils), apoptosis, exudatesclot formation could not be utilized. Instead, areas of tissue injury were identified by quantifiable denudation of the normal layers of the intestine or from increased thinning and elongation of the cell with dense hyperchromatic nuclei. Two metrics of damage were quantified: an intestinal layer thickness calculation where the serosa (outermost) layer was measured in the area of compression (C) and compared to a local control (LC) region that was not compressed as a percent deformation and a histological scoring scale for tissue trauma (Figure 2). The histological scoring scale was created by the two pathologists in this study (and employed in our previous preliminary study), as we were unable to find a suitable pathologist-validated scale endorsed in the literature [23]. The criteria for the scale is outlined in Table 1 and representative images shown in Figure 2.

**TABLE 1.** Tissue TraumaScoring Criteria

Tissue Stress from Graspers and Bowel Injury
| *Trauma Score* | *Cellular Architecture* |
| --- | --- |
| Grade 0<br>(no trauma to the serosa) | Nuclei of serosal and bounding muscularis externa cells are smooth, oval and uniformly shaped. |
| Grade 1<br>(minor trauma to the serosa) | Elongation and mild hyperchromasia of nuclei in muscle/connective tissue in both the serosa and muscularis propria externa outer longitudinal layer, but in less than 50% of cells in our region of interest (ROI), representing trauma to the cells. |
| Grade 2<br>(significant trauma to the serosa) | Clear and significant damage to the serosa and muscularis propria externa outer longitudinal layer, with more than 50% of nuclei in the cells in our ROI appearing significantly elongated and thinned and there is the presence of multiple hyperchromatic nuclei. |
| Grade 3<br>(complete denudation of the serosa) | The serosa and muscularis externa longitudinal layers are both compressed, with evidence of denudation of the serosa and trauma extending to the muscularis propria inner circular layer. |

Histological images were taken at 400x and all serosa layer thickness measurements were performed at the center of the experimental area of compression (as visualized on the scanned slides as areas where dye was present from the indentation tip) as the center region is the most representative of the average stress of the transmitted force.

Five measurements were taken of the thickness of the serosa layer in the area of compression and then averaged for a total value. Five measurements were also taken in an adjacent non-compressed area and averaged to serve as a local control. Multiple data points were sampled for each patient to counteract artifacts associated with the typical histological slide creation process. This includes tissue shrinkage or the warping of tissue orientation on slides. Percent deformation (rather than an absolute delta in microns) was used for this reason as well, and was calculated as the percent difference between the thicknesses of the compressed area to its local control of uncompressed tissue.

### Experimental Methodology

The PrecisionCAT was created to deliver a precise and discrete pressure “grasp” to the loaded tissue sample via compression by the two grasp plates to simulate being grasped by a laparoscopic grasper. As previously discussed above, we chose to program loading conditions of 0 to 11.8 N or 0 to 600 kPa (specifically of 2 N, 3.9 N, 5.9 N, 7.8 N, 9.8 N and 11.8 N or using our precision pin plate with known indentation tip surface area of 19.6mm^2^, calculated pressures of 0 kPa, 100 kPa, 200 kPa, 300 kPa, 400 kPa, 500 kPa and 600 kPa) for a duration of 10 seconds to match those previously reported in porcine literature. Logically, we would expect to induce more tissue trauma as we apply higher loads, with the most significant damage occurring at the highest loads of 9.8 N (500 kPa) and 11.8 N (600 kPa) of pressure. A new piece of 1 cm x 1 cm tissue was used for each loading condition and was compressed at a loading rate of 5 mm/s until making initial contact with the tissue (as defined by a positive mean force of the last three measurements) at which point velocity slowed to 1 mm/s. The velocity was programmed to progressively reduce by half as force approached the target force in order to maintain a minimum force resolution of approximately 1% of the target force. This asymptotic approach for velocity is necessary to minimize overshoot due to the rapid increase in tissue stiffness as it is compressed. Once the target force was achieved, the system then maintained this position for a duration of 10 seconds, which represents a typical duration of grasp during laparoscopic surgery, before releasing the tissue at the same rate as it was loaded. The signal voltage was filtered from the force transducer (load cell) using a low pass filter (a -3dB Bessel filter at 2 kHz) to smooth out unwanted noise. Position, time and force values were all logged synchronously at 31 Hz and stress and strain were calculated (Figure 3). During statistical analysis, force data was further pre-processed using a third-order Butterworth low-pass digital filter with a cut-off frequency of 12.4 Hz. Mechanical measures of stress and strain were calculated from the raw positional and force data.

### Statistical Analysis

Statistical analysis and modelling was performed using NumPy, pandas and Scikit-learn using the Python programming language [32]–[34]. All graphs were created with Matplotlib [35]. A series of one-tailed t-tests were performed to compare sample serosal thickness measurements at compressed vs local control area in the histological slides. The tests were to investigate significant decrease in thickness due to compression of the tissue. Different groupings were considered in the analysis by patient, load level and tissue type. To investigate the relationship between serosal thickness change and the pathologist’s trauma score ratings, a correlation study was also completed. The goal of this analysis was to establish evidence of tissue damage and to identify promising predictive metrics that could be utilized in a live operative environment. As such, features derived from the histological analysis would be unable to be used in real time, and were excluded for the predictive models.

A logistic regression model was trained to predict significant serosal thickness change as a classification task. A second logistic regression model was trained to predict a positive tissue trauma score. The intention of these predictive models was evaluate how well input features that could be measured intraoperatively such as force, position of the grasper and time that the tissue was compressed and features that could be derived from those such as stress, strain and stiffness of the tissue, could be used to predict the likelihood of subsequent tissue trauma. This algorithm then could be incorporated into a “smart” laparoscopic tool that can, in real-time, alert a surgeon in the operating room about force use and subsequent pathological tissue response.

## Results

### Demographics and Experimental Cases

The research team was limited in regard to the availability of specimens, as all tissue used in this study were from scheduled operations and were tissues taken out as per normal surgical workflow. Overall, 26 samples were procured but six tissue samples were unusable due to a variety of factors such as the pathologists being unavailable for immediate experimentation, the tissue being too inflamed, or the tissue sample being of inadequate size to complete the experimental protocol. In total, we were able to complete the full experimental protocol (6 load pressures) on 20 patients’ tissues, with 10 colon samples and 10 small bowel samples for a total of 120 experimental cases and 120 control cases (Table 2).

**TABLE 2.** Patient Demographics

| *Tissue type (n)* | *Gender (M:F)* | *Age (Mean±SD, Years)* | *Surgical Procedure* |
| --- | --- | --- | --- |
| Small Bowel (10) | 7:3 | 54.3 ± 18.17 | Ileocolic resection (5)<br>Loop ileostomy (2)<br>Small bowel resection (3) |
| Colon (10) | 1:1 | 65.2 ± 17.96 | Loop ileostomy (1)<br>Right hemicolectomy (2)<br>Rectal prolapse (1)<br>Sigmoid colectomy (2)<br>Anterior resection (3)<br>Abdominal perineal resection (1) |

### Tissue measurements

#### i. Slide creation and artifact rate

As previously mentioned, the process of creating histological slides from tissue that has been subjected to a load force has an associated failure rate, which we have taken numerous steps from our previous study to optimize. The following table (Table 3) demonstrates the loss we experienced in the amount of useable histological slides prepared. Out of a possible 120 experimental tissue slides, a final useable total of 104 cases (86.7%) were obtained.

**TABLE 3.**
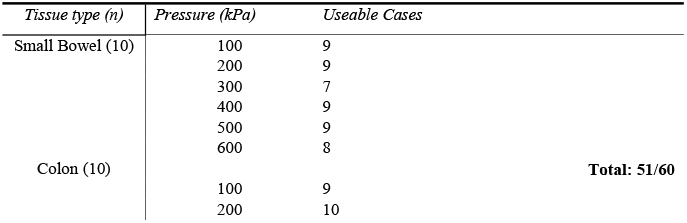
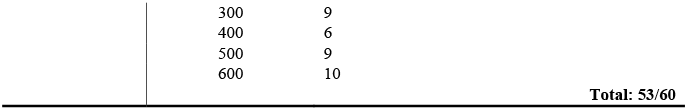
Slide creation and Artifact Rate

| *Tissue type (n)* | *Pressure (kPa)* | *Useable Cases* |
| --- | --- | --- |
| Small Bowel (10) | 100 | 9 |
|  | 200 | 9 |
|  | 300 | 7 |
|  | 400 | 9 |
|  | 500 | 9 |
|  | 600 | 8 |
| Colon (10) |  | **Total: 51/60** |
|  | 100 | 9 |
|  | 200 | 10 |
|  | 300 | 9 |
|  | 400 | 6 |
|  | 500 | 9 |
|  | 600 | 10 |
|  |  | **Total: 53/60** |

#### ii. Average serosa layer thickness

Average serosa thickness of both the small bowel and colon was calculated by averaging each tissue type’s local control area measurements (Table 4, excluding slides that had unsatisfactory histological slide creation). These values fall within range of those reported in the literature [36], [37].

**TABLE 4.**
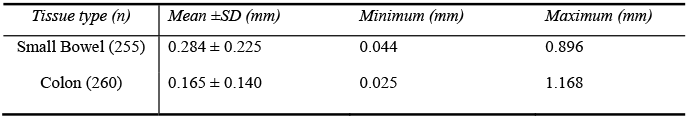
Serosa thickness Analysis

| *Tissue type (n)* | *Mean ±SD (mm)* | *Minimum (mm)* | *Maximum (mm)* |
| --- | --- | --- | --- |
| Small Bowel (255) | 0.284 ± 0.225 | 0.044 | 0.896 |
| Colon (260) | 0.165 ± 0.140 | 0.025 | 1.168 |

#### iii.Serosa thickness deformation

A series of paired t-tests were conducted between the five serosal control measurements and the five compression site measurements at each loading condition to determine significance (Table 5), where p-values ≤0.05 are indicated by bold text and missing values indicate that the slide was unable to be analyzed due to slide artifacts.

All 20 patients experienced a significant difference (p<0.05) in serosal thickness post-compression at both 500 and 600 kPa for both tissue types. The majority of patients at 400 kPa (6/10 for small bowel and 5/6 for colon) also had significant differences pre- and post-compression. This is similar to our pilot study where all patients had significant p-values starting at 450 kPa [23]. This is also higher than the minimum pressure of 150 kPa that Chandler et al.’s group found for damage to porcine colon, but their measurements were for the mucosal and muscle layers of the colon and did not focus on serosal change as they had this layer of tissue stripped [24]. There were no significant changes at 100 and 200 kPa (except for patient 20 for colon) and a mixed picture at 300 kPa with 50% (8/16) of patients experiencing a significant difference between control and compressed tissues (4/7 for small bowel and 4/9 for colon). This data is visually displayed in Figure 3b and c. When all small bowel patient data was combined together and analyzed by loading condition, the trend was similar to individual results. Significance is achieved at 300 kPa and continues onwards from 400 – 600 kPa with significant p-values. This is similar to grouped colon patient data, where significance is achieved at 400 kPa and continues onwards to 600 kPa with significant p-values. All patient data was then grouped together (both small bowel and colon) and analyzed by loading condition and this too followed a comparable pattern where significance is achieved at 300 kPa and continues onwards from 400 – 600 kPa with p-values of <0.001. Serosal thickness as a percent deformation (to normalize measurements) was also calculated for each patient and plotted in Figure 4a. Percent deformation steadily increases as the experimental loading condition is increased and both tissue types follow similar patterns (Figure 4b, c).

**TABLE 5.**
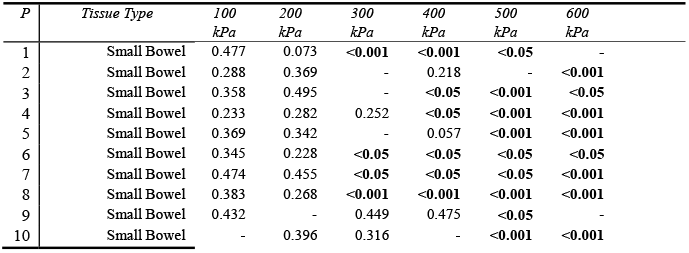
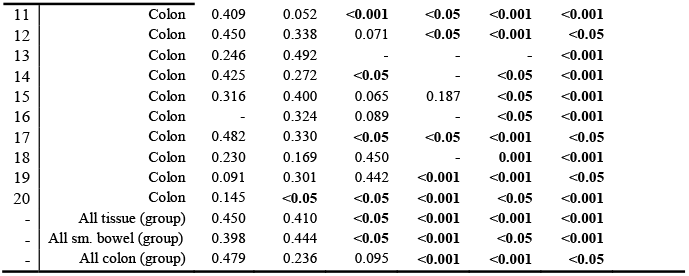
T-test results Between Control andCompression Sites

| *P* | *Tissue Type* | *100*<br>*kPa* | *200*<br>*kPa* | *300*<br>*kPa* | *400*<br>*kPa* | *500*<br>*kPa* | *600*<br>*kPa* |
| --- | --- | --- | --- | --- | --- | --- | --- |
| 1 | Small Bowel | 0.477 | 0.073 | **<0.001** | **<0.001** | **<0.05** | - |
| 2 | Small Bowel | 0.288 | 0.369 | - | 0.218 | - | **<0.001** |
| 3 | Small Bowel | 0.358 | 0.495 | - | **<0.05** | **<0.001** | **<0.05** |
| 4 | Small Bowel | 0.233 | 0.282 | 0.252 | **<0.05** | **<0.001** | **<0.001** |
| 5 | Small Bowel | 0.369 | 0.342 | - | 0.057 | **<0.001** | **<0.001** |
| 6 | Small Bowel | 0.345 | 0.228 | **<0.05** | **<0.05** | **<0.05** | **<0.05** |
| 7 | Small Bowel | 0.474 | 0.455 | **<0.05** | **<0.05** | **<0.05** | **<0.001** |
| 8 | Small Bowel | 0.383 | 0.268 | **<0.001** | **<0.001** | **<0.001** | **<0.001** |
| 9 | Small Bowel | 0.432 | - | 0.449 | 0.475 | **<0.05** | - |
| 10 | Small Bowel | - | 0.396 | 0.316 | - | **<0.001** | **<0.001** |
| 11 | Colon | 0.409 | 0.052 | **<0.001** | **<0.05** | **<0.001** | **<0.001** |
| 12 | Colon | 0.450 | 0.338 | 0.071 | **<0.05** | **<0.001** | **<0.05** |
| 13 | Colon | 0.246 | 0.492 | - | - | - | **<0.001** |
| 14 | Colon | 0.425 | 0.272 | **<0.05** | - | **<0.05** | **<0.001** |
| 15 | Colon | 0.316 | 0.400 | 0.065 | 0.187 | **<0.05** | **<0.001** |
| 16 | Colon | - | 0.324 | 0.089 | - | **<0.05** | **<0.001** |
| 17 | Colon | 0.482 | 0.330 | **<0.05** | **<0.05** | **<0.001** | **<0.05** |
| 18 | Colon | 0.230 | 0.169 | 0.450 | - | **0.001** | **<0.001** |
| 19 | Colon | 0.091 | 0.301 | 0.442 | **<0.001** | **<0.001** | **<0.05** |
| 20 | Colon | 0.145 | **<0.05** | **<0.05** | **<0.001** | **<0.05** | **<0.001** |
| - | All tissue (group) | 0.450 | 0.410 | **<0.05** | **<0.001** | **<0.001** | **<0.001** |
| - | All sm. bowel (group) | 0.398 | 0.444 | **<0.05** | **<0.001** | **<0.05** | **<0.001** |
| - | All colon (group) | 0.479 | 0.236 | 0.095 | **<0.001** | **<0.001** | **<0.05** |

#### iv.Tissue trauma score

Tissue trauma scores were assigned by both pathologists for each histological slide created based on the criteria outlined in Table 1 (plotted in Figure 4d). For small bowel (Figure 4e), at 600 kPa, all 8 patients experienced trauma to the serosa with 1/8 patients graded as a Trauma Score of 2, and 7/8 patients being graded as a Trauma Score of 1. At 500 kPa, 8/9 patients were graded with a Trauma Score of 1 with the remaining patients graded as no trauma. At 400 kPa, the picture is mixed as 5/9 patients’ tissues were graded with a Trauma Score of 1 and the remaining 4 patients displaying no trauma. At 300 kPa, 3/7 patients were graded with a Trauma Score of 1 while 4/7 exhibited no trauma. At 200 kPa only 2/9 patients were graded with a Trauma Score of 1 and 7/9 patients had no evidence of tissue trauma. Lastly, at 100 kPa, no patient had a positive Trauma Score.

For colon (Figure 4f), at 600 kPa, all 10 patients experienced trauma to the serosa with 2/10 patients graded as a Trauma Score of 2, and 8/10 patients being graded as a Trauma Score of 1. At 500 kPa, all 9 patients again had positive scores with 2/9 patients graded with a Trauma Score of 2 with the remaining 7 patients graded with a Trauma Score of 1. At 400 kPa, again all patients were rated as displaying evidence of trauma with all 6 patients receiving a score of 1. At 300 kPa the picture is mixed, with 6/9 patients graded with a Trauma Score of 1 and the remaining 3 displaying no tissue damage. At 200 kPa, the majority (7/10) of patients were rated with a score of 0 and 3/10 patients graded with a score of 1. Lastly, at 100 kPa, again, the majority of patients (6/9) were scored as displaying no trauma and 3/9 being graded with a Trauma Score of 1.

Out of 104 experimental samples, 83 samples (79.8%) were classified in agreement between the two available metrics: Trauma Score and significant serosa thickness change (Figure 5). This means they were either both classified as having a positive Trauma Score and having significant serosa deformation or were both classified as having a Trauma Score of 0 and non-significant serosa deformation. Of the samples with disagreeing classification, 7/104 (6.7%) showed significant change in serosa thickness but were assigned a Trauma Score of 0 and 14/104 (13.5%) were assigned a Trauma Score of 1 or greater but did not display evidence of significant serosa thickness change.

### Logistic Regression Model

Two logistic regression models were trained in order to predict target metrics: a positive Trauma Score and significant serosa thickness change. Logistic regression was selected because it is an easily interpretable model and could be implemented in an intraoperative environment in a manual or automatic approach. Other models (especially Support Vector Machine classifiers) were able to outperform the logistic regression in predictive trials on the data. However, logistic regression will be considered in this paper because of its ease of implementation in a real-time setting. In addition, the models were trained on a single feature (target stress, Figure 6a), which was selected as it was the most predictive (it was the feature with the highest correlation). This feature was expected to be the most important to predict tissue trauma because it is directly derived from the maximum force.

The features that both models included were derived from the force and displacement measurements obtained during each experimental condition. Firstly, the force measurements were transformed to stress on the tissue based on the contact surface area of the indentation pin. The displacement measurements, measured with a resolution of 1 µm, were converted into strain of the tissue, taking into account the initial tissue thickness, as determined by the tool coming into contact with the tissue. Both of these measurements, in addition to time stamps, could be measured in a sensorized grasper tool.

From this data, further features were derived, which is shown in Table 6. Additionally, patient age, gender and tissue type were included as features as well.

**TABLE 6.**
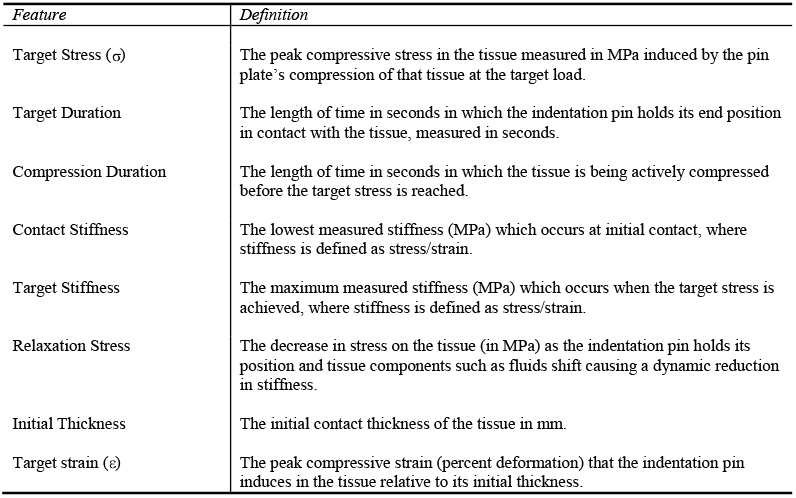
Logistic RegressionModel Features

| *Feature* | *Definition* |
| --- | --- |
| Target Stress ($\sigma$) | The peak compressive stress in the tissue measured in MPa induced by the pin plate's compression of that tissue at the target load. |
| Target Duration | The length of time in seconds in which the indentation pin holds its end position in contact with the tissue, measured in seconds. |
| Compression Duration | The length of time in seconds in which the tissue is being actively compressed before the target stress is reached. |
| Contact Stiffness | The lowest measured stiffness (MPa) which occurs at initial contact, where stiffness is defined as stress/strain. |
| Target Stiffness | The maximum measured stiffness (MPa) which occurs when the target stress is achieved, where stiffness is defined as stress/strain. |
| Relaxation Stress | The decrease in stress on the tissue (in MPa) as the indentation pin holds its position and tissue components such as fluids shift causing a dynamic reduction in stiffness. |
| Initial Thickness | The initial contact thickness of the tissue in mm. |
| Target strain ($\epsilon$) | The peak compressive strain (percent deformation) that the indentation pin induces in the tissue relative to its initial thickness. |

#### i. Serosa Deformation Model

The first model (Figure 6b) to predict significant serosa deformation uses all the features derived from the force, position and time measurement data. It is able to achieve a sensitivity of 100% and a specificity of 84.6% on a test set of data (20% of total cases). The receiver operating characteristic (ROC) curve (Figure 6b) shows the performance of the classification model at all thresholds and can be used to assess its performance as compared to random guessing. The area under the curve (AUC) is 92%.

The model was retrained using only target stress as an input feature. The advantage of this restricted approach is that it only needs intraoperative force measurements which potentially can simplify what sensors are needed in a sensorized grasper. The performance of the model (Figure 6c) deteriorated relative to the full featured model, with sensitivity dropping to 75% and specificity remaining the same at 84.6%. The AUC dropped to 80%.

#### ii. Trauma Score Model

The first model (Figure 6d) to predict positive Trauma Score using all the features, is able to achieve a sensitivity of 76.9% and a specificity of 100% on a test set of data (20% of total cases). The area under the curve (AUC) is 88%.

The model was retrained using only target stress as an input feature. The performance of the model (Figure 6e) deteriorated relative to the full featured model, with sensitivity dropping to 61.5% and specificity remaining the same at 100%. The AUC dropped to 81%.

#### iii. Model Analysis

The first metric of significant serosa deformation with all features has a high sensitivity and thus will identify all cases of trauma successfully but may have some false positives. The second metric of positive Trauma Score has a high specificity and thus will identify patients who do not have significant trauma but may have some false negatives. There may be value in applying both models as conservative prediction is preferred.

Both models showed a deterioration when features were restricted to target stress only. There may be advantages in implementation however in only measuring intraoperative forces with sensorized tools, so this degradation in performance is presented as a comparison. The suitability of these restricted models would need to be evaluated in a clinical setting.

Other models were identified that showed higher prediction accuracy on the data set such as Support Vector Machines, however they have a higher computational complexity for implementation than the logistic regression models presented here.

#### iv. Clinical Thresholds

For clinical application, there is a benefit in a single feature predictive model, in that it can provide a hard threshold, above which, tissue trauma is likely. Both restricted models that focus on target stress only for both metrics agree on this threshold stress value within 1 kPa. The significant serosa deformation model predicts a threshold for damage at 330.3 kPa and the Trauma Score model predicts a threshold of 329.3 kPa. Thus, we propose that the use of bowel graspers should not exceed this value of compressive stress when handling bowel tissues.

## Discussion

This paper aimed to build upon our previous work with a more sophisticated compressive device that had the additional capabilities of force, time and position logging. We were also able to optimize both our histology slide preparation protocol and patient recruitment to run a more comprehensive study. Despite the importance of elucidating the relationship between compressive force and tissue trauma, only a few studies have been performed to date. As far as we know, we are the first group to publish a paper based on compressive data of human gastrointestinal tissues via a histological analysis with such a large amount of patient tissue data. The histological analysis was central to this study as it allowed us to objectively identify at which loading conditions mechanical trauma occurred.

What needs to be elucidated further in human studies, is if trauma occurs to a tissue, at what point is that tissue’s trauma pathological? Our data shows significant serosal thickness change occurring at 300 kPa for both the small intestine and colon, which also correlated to a marked increase in Trauma Score by the pathologists. What we don’t know is if the body would have the ability to recover from the damage we caused or whether this force level induces dysfunctional healing. What we do know from Heijnsdijk’s work with porcine and human bowel tissues is that the inter-individual variability in perforation forces is large and that bowel strength could differ by a factor of two between patients [20]. Taking this into account we must be extremely careful and conservative when defining “safe” force limits for intestinal tissue because a force that could be safely applied to one patient may cause a perforation in another patient.

Using our logistic regression analysis, our data points in the direction of establishing a maximum force cut-off starting at 329 kPa on average for gastrointestinal tissues using a 50% threshold, however very large safety margins should be considered and used. This is similar to the results of our pilot study which suggested a damage threshold of 350 kPa for colon tissues [23]. These results need to be viewed in context of the limitations of an *ex-vivo, in-vitro* study, with the chief caveat revolving around the inability to see how these forces affect tissue *in-vivo*. How much serosal damage can the body repair and at what point does the repair mechanism become dysfunctional and leads to adhesion formation or necrosis needs to be further elucidated in a longitudinal, *in-vivo* study that can quantify objective markers of inflammation and cellular death. Intestinal force cut-offs also need to be determined for pathological tissues (such as inflamed colon in Crohn’s disease) to ensure intraoperative tissue handling is accounted for in both healthy and diseased states. Our study was also performed on bowel tissue that we rinsed clean before experimentation, but intraoperatively, if a patient were not subject to bowel preparation, bowel content would lead to the bowel being more distended and therefore more susceptible to trauma.

In context of the current existing literature, Heijnsdijk’s study saw perforation of the human small bowel at 10.3±2.9 N of force, but that was using a sharp pinch device with a smaller diameter than our pin plate (thus generating higher pressure loads), with weights that were manually loaded and moved by the experimenter. Culmer et al.’s team showed trauma occurring at 150 kPa but that was in a porcine model, and porcine tissues have a lower ultimate tensile strength and elastic modulus than human tissues do [19]. What our study makes clear and confirms from these two previous studies is that tissue damage definitively correlates to how much force is exerted on that tissue in compression. The two pathologists who were blinded to the loading condition of the tissue were both independently able to quantify increasing damage amounts to increasing pressure; this relationship was true regardless of the tissue type. Our study is a foundational study that other groups may build upon using our standardized methodology. We have released the entire build of materials, schematics, 3D-printing files and Python code necessary to replicate this study and invite research groups that would like to reproduce our study to do so, as multisite validation with a much larger *n* number than we could produce is necessary to authenticate our results. While this study focused on the testing of small and large intestine (due to the significance of an intraoperative injury with laparoscopic graspers), these methods could be applied to other delicate tissues of interest such as the ureter, bile duct and fallopian tubes as well.

Our paper adds strong evidence for the practical use of this information, namely, that if an upper limit of atraumatic force can be reliably established in humans, this force cut-off should be used intraoperatively via laparoscopic graspers “smart” tools. The two logistic regression equations created could be utilized in a tool intraoperatively, provided that the features used could be measured in real-time. This would require a minimum of two sensors: one for force and one for grasper jaw position, similar to the PrecisionCAT measure *in-vitro*. Sensorized laparoscopic graspers can provide real-time force information to an operating surgeon, who can then limit themselves to this “safe zone”. Of course force limits would be tissue-type specific as a tissue’s ability to handle pressure is a function of its underlying cellular structure. However, the outcomes of this study are a promising step forward in the field of tissue trauma prediction and prevention in surgery. Future work will focus on increasing our patient recruitment so that we can have a higher *n* number to power our histological slide analysis.

## Data Availability

All files and schematics used to create the PrecisionCAT is freely available on the Harvard Dataverse Repository.
All custom Python code created and used in the preparation of this manuscript is freely available on our Github.

https://dataverse.harvard.edu/dataset.xhtml?persistentId=doi:10.7910/DVN/Q0WOEZ

https://github.com/crushdevice

## Acknowledgements

We would like to first thank all of the patients who participated in our study and graciously allowed us the use of their tissues for the advancement of science. Second, the Hospital for Sick Children’s Perioperative Services Grant kindly funded the creation of the PrecisionCAT in collaboration with the Centre for Image-Guided Innovation and Therapeutic Intervention (CIGITI). We also extend our gratitude to the general surgeons from St. Michael’s Hospital in Toronto who allowed us into their operating rooms and Dr. Shuning “Steve” Bian for allowing us to adapt his Python code. We are also in debt to Drs. Peter Culmer and James Chandler from the School of Mechanical Engineering at the University of Leeds who created the original crush apparatus that inspired our design. We would also like to thank Drs. Anne Agur, Ted Gerstle, Joel Krivy and Hani Naguib for their wisdom, brainstorming and feedback, which this paper would not be published without. Lastly, we would like to thank our long-time collaborator Granitus MKM for her always useful input, proofreading and commentary on the manuscript. Portions of this manuscript have been previously published as part of AFK’s PhD dissertation.

## Data Availability

All files and schematics used to create the PrecisionCAT is freely available on Harvard’s Dataverse Repository at the following URL: https://dataverse.harvard.edu/dataset.xhtml?persistentId=doi:10.7910/DVN/Q0WOEZ

## Code Availability

All custom Python code created and used in the preparation of this manuscript is freely available on Github. The PrecisionCAT data is available, freely to the public at the following URL: https://github.com/crushdevice

## Build of Materials

| Part | Description | Product Code |
| --- | --- | --- |
| SMAC Linear Actuator | 25 mm stroke; 48 volts | https://www.smac-mca.com/lca-series-p-15.html?cPath=1_4<br>LCA 50-025-72-1F-3 |
| Power Supply | 48 volt | Generic |
| Omega Load Cell | Miniature universal load cell with $\pm 5$ kgF range | https://www.omega.com/pptst/LCM703.html<br>LCM703-5 |
| LAC-1 Controller | Single axis controller; 12-48 volts | https://www.smac-mca.com/lac-single-axis-controller-p-127.html |
| LAC-1 Communication Kit | Used for interfacing with the controller | Bundled with LAC-1 Controller |
| Omega Load Cell Strain Gage Amplifier | Bridge excitation of 4 to 15 volts; adjustable gain and offset | https://www.omega.ca/en/communication-and-connectivity/signal-conditioners-and-transmitters/signal-conditioners/dmd-465-series/p/DMD-465<br>DMD-465 |
| End-Feed Fastener | M6 thread for 30 mm high rail T-slotted framing | https://www.mcmaster.com/5537t527<br>McMaster-Carr part 5537T527 |
| T-Slotted Framing Single Rail | 1 ft, 30 mm x 30 mm | https://www.mcmaster.com/5537t97<br>McMaster-Carr part 5537T97 |
| T-Slotted Framing Corner Brackets | For 30 mm | https://www.mcmaster.com/5537t936<br>McMaster-Carr part 5537T936 |
| 316 Stainless Steel Threaded Rod (20 mm) | M6; 1 mm x 20 mm | https://www.mcmaster.com/98863a243<br>McMaster-Carr part 98863A243 |
| 316 Stainless Steel Threaded Rod (40 mm) | M6; 1 mm x 40 mm | https://www.mcmaster.com/98863a260<br>McMaster-Carr part 98863A260 |
| 316 Stainless Steel Thin Hex Nut (M6) | M6; 1 mm | https://www.mcmaster.com/93935a335<br>McMaster-Carr part 93935A335 |
| 316 Stainless Steel Thin Hex Nut (M3) | M3; 0.5 mm | https://www.mcmaster.com/93935a320<br>McMaster-Carr part 93935A320 |
| Aluminum Plate | 6061 aluminum | Custom milled |

---

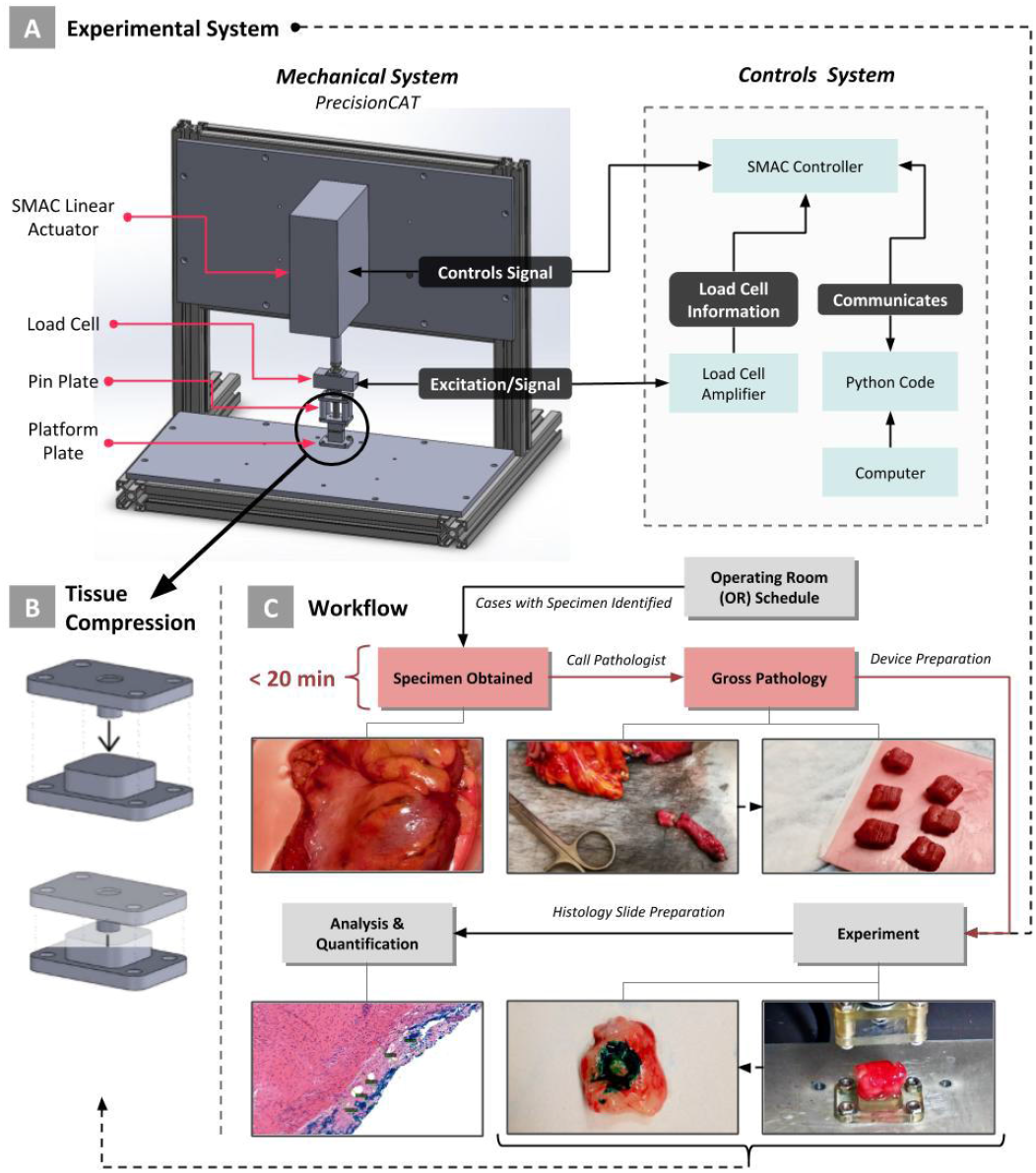

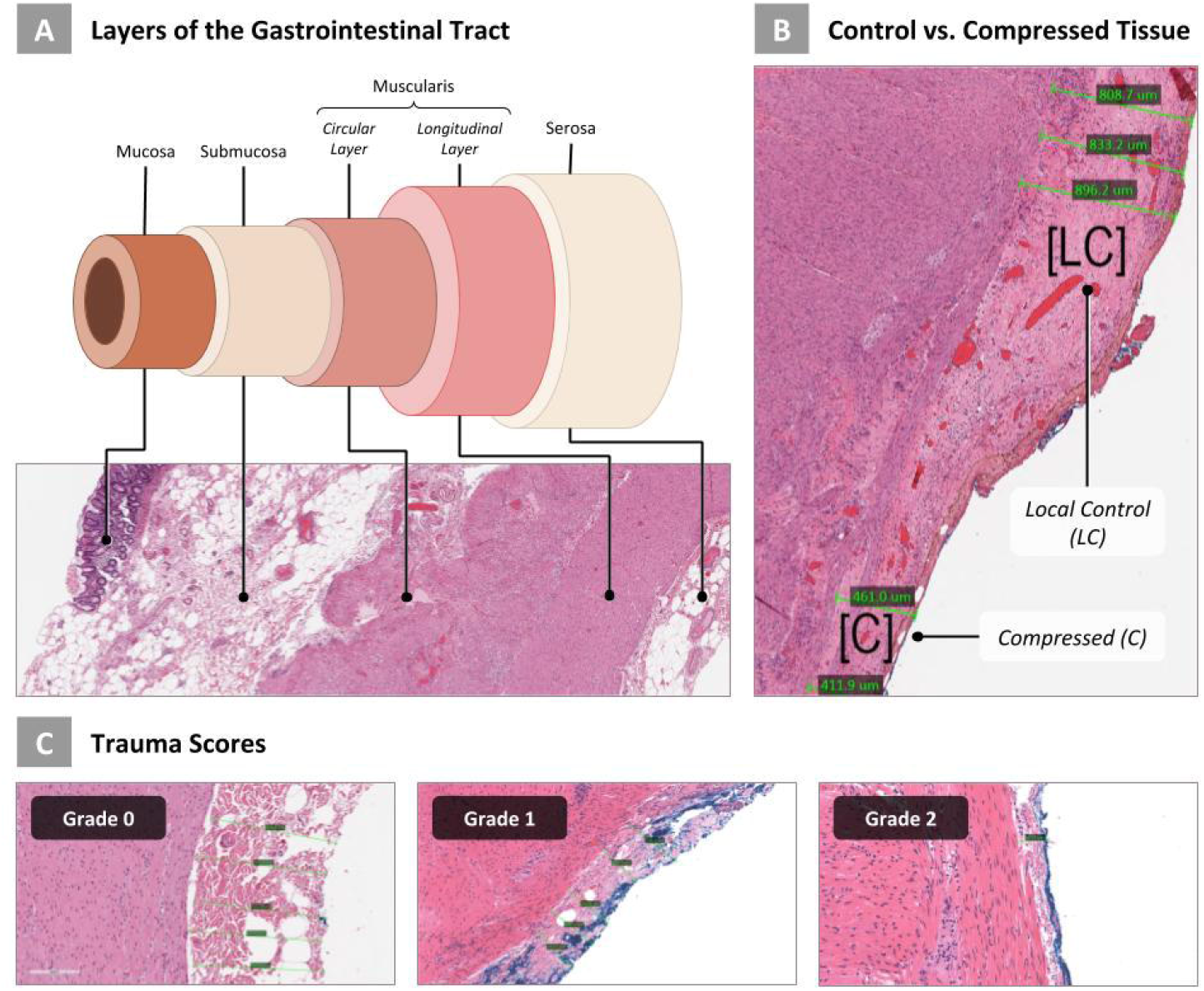

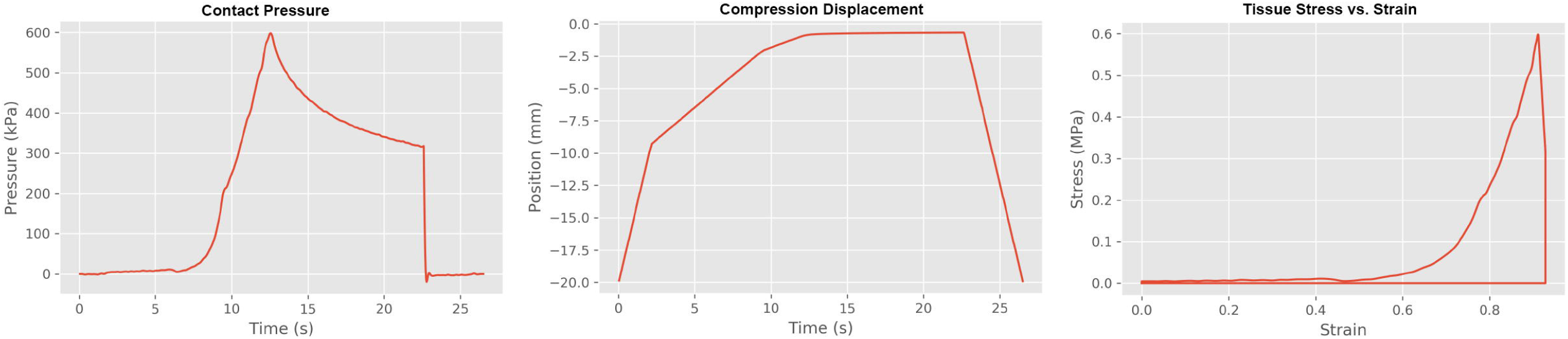

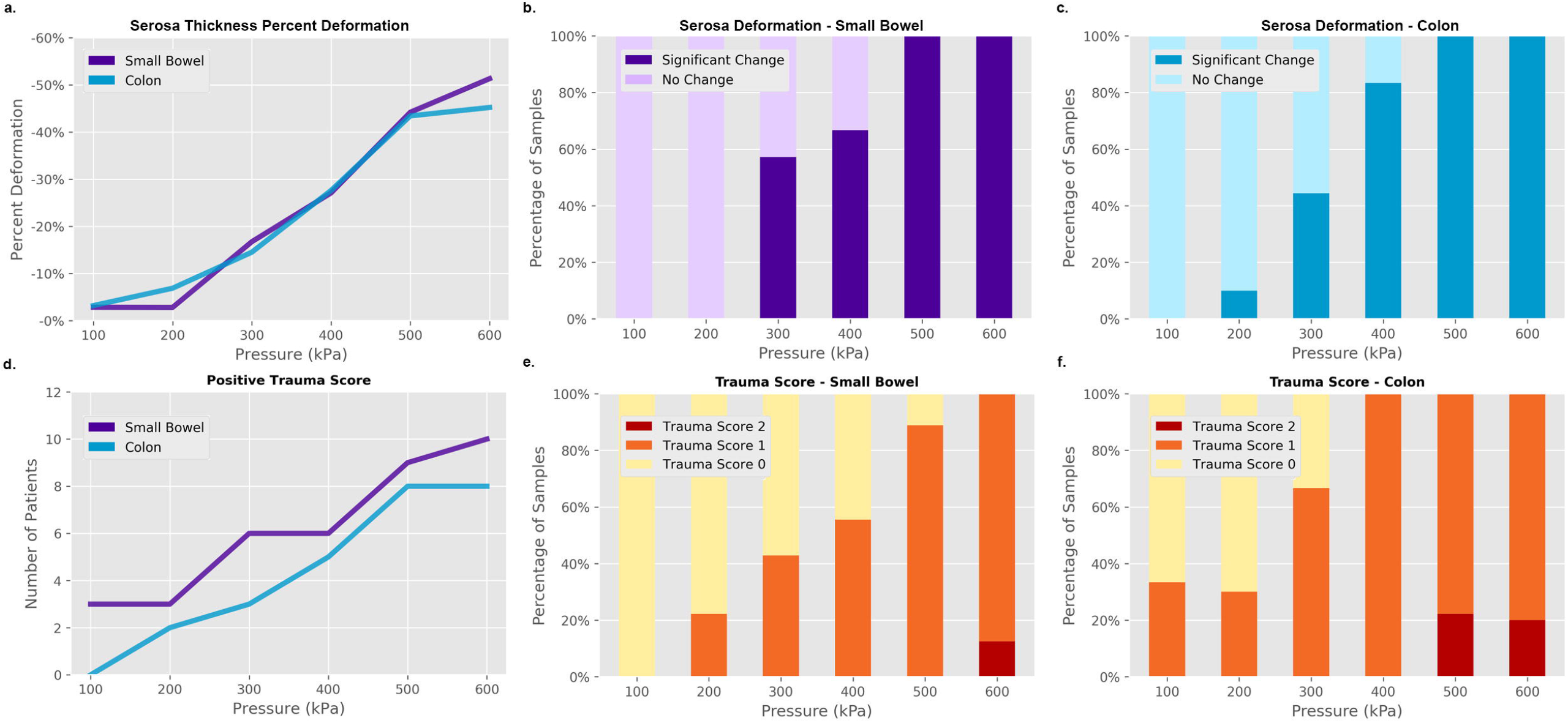

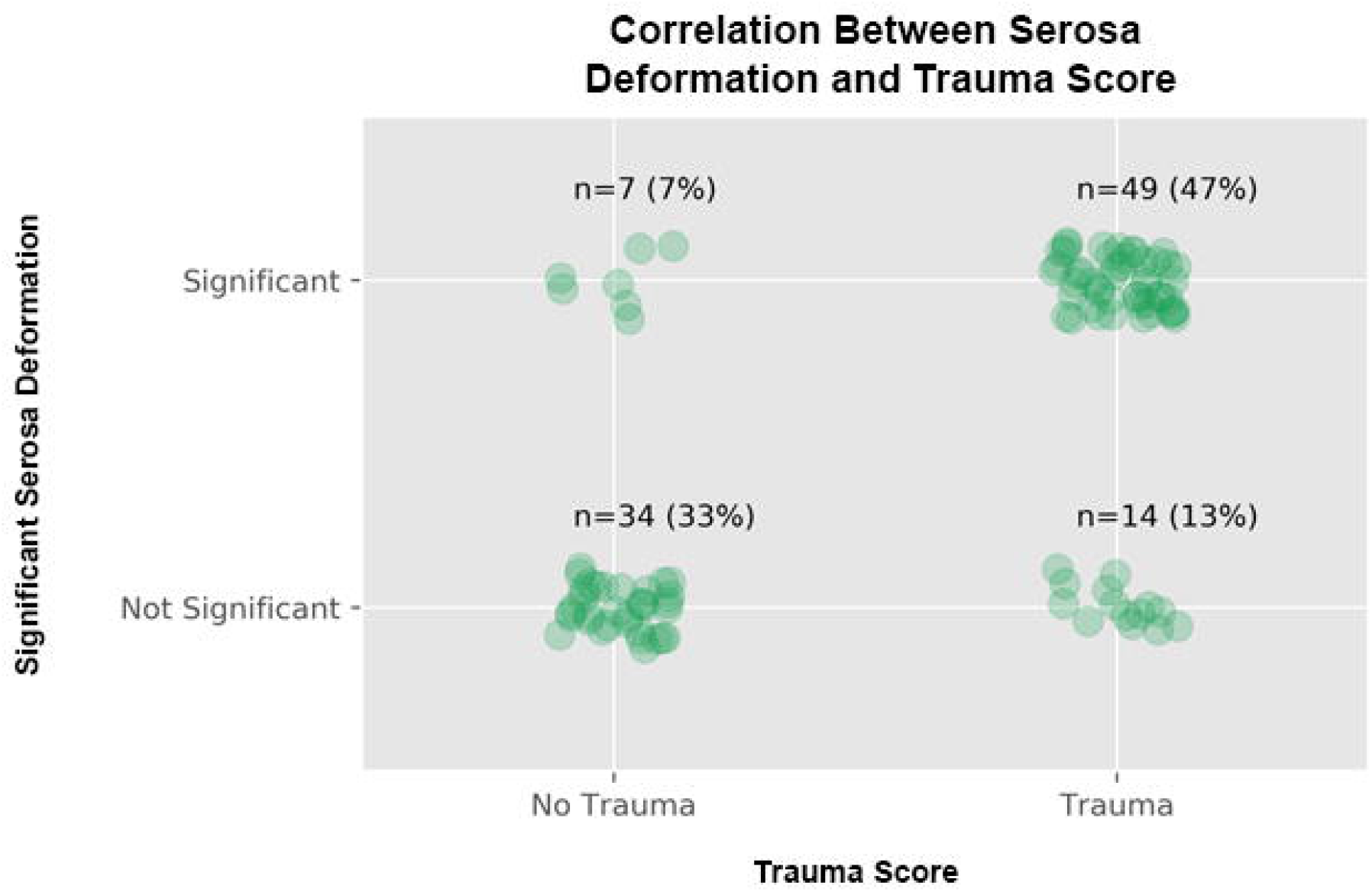

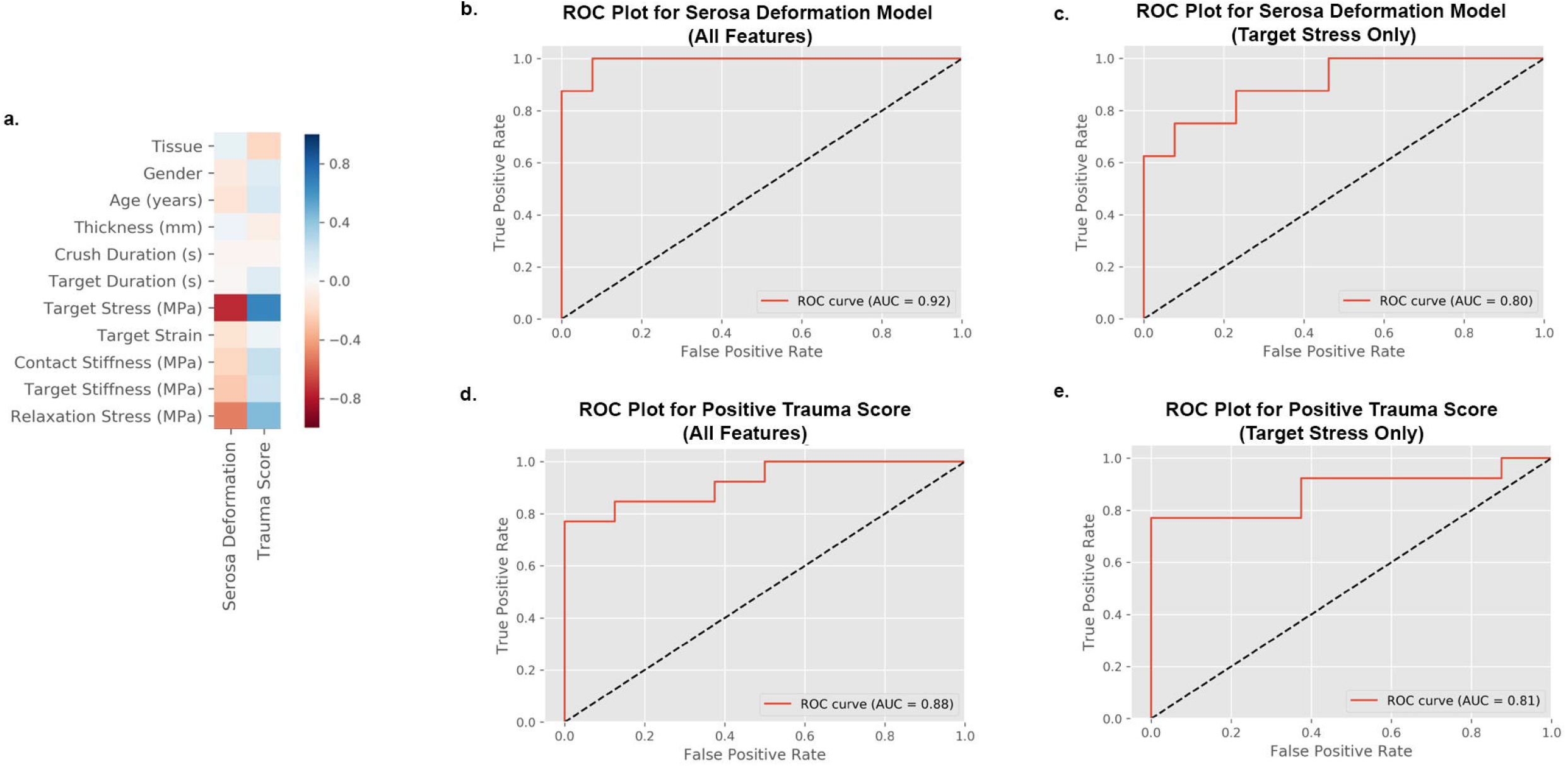

